# Identifying Social-Epidemiological Roles Associated with Viral Exposure Using Regular Equivalence Blockmodeling

**DOI:** 10.1101/2025.11.05.25339611

**Authors:** Tyler M. Barrett, Charles Kevin Tiu, Kayla Kauffman, Michelle Pender, Jean Yves Rabezara, Prisca Rahary, Lin-Fa Wang, Randall A. Kramer, Voahangy Soarimalala, Peter J. Mucha, James Moody, Charles L. Nunn

## Abstract

Identifying individuals at high risk of infection is critical to guiding interventions during infectious disease outbreaks. Network centrality can characterize infection risk, but its utility varies across pathogens and social systems. We used equivalence-based blockmodeling to identify social-epidemiological roles associated with viral exposure among 1,297 adults in northeast Madagascar. Roles were determined based on social networks derived from reported shared free time and exchanges of food and farmwork. We identified three distinct role categories, including individuals with many reciprocated social ties (Popular), individuals who sent many ties with few reciprocated (Hangers-On), and individuals who had few connections (Periphery). To assess whether the role categories covaried with viral exposure, we performed Phage ImmunoPrecipitation Sequencing with VirScan using dried blood spot samples to determine exposure to 337 virus species and subtypes. Individuals in the Popular (*ß* [95% CI]: 0.29 [−0.06-0.63]) and Hangers-On (*ß* [95% CI]: 0.36 [0.12-0.60]) role categories had greater viral exposure than individuals in the Periphery role category. Roles performed better at predicting exposure than single measures of centrality. Equivalence-based blockmodeling extends the utility of network centrality for characterizing infection risk and provides new insights into how social roles relate to both pathogen exposure and susceptibility to infection.

## Introduction

The social patterning of infectious disease depends on *whom* you interact with and *how* you interact with them. Advances in network analysis have highlighted the importance of this individual-level heterogeneity for epidemic dynamics. For example, superspreading – where individuals disproportionately infect many other individuals in a population – helped drive the explosive spread of SARS-CoV-1 during the 2002-2004 pandemic [1, 2]. Superspreading has also been important for understanding the transmission of SARS-CoV-2, Ebola, and measles [3–5]. Network centrality-based approaches have since been used to identify individuals with superspreading potential and high risk of infection, i.e., individuals who have many connections (e.g., degree centrality) or specific types of connections (e.g., betweenness centrality) that are known to facilitate parasite transmission [6–12].

Simulation-based studies have shown that an individual’s risk of infection and time to infection over the course of a simulated epidemic are associated with multiple measures of centrality, including degree, strength, betweenness, and eigenvector centrality [6, 7, 13–18]. Observational studies of infection patterns across a diverse range of host and parasite species also revealed that more central individuals have a higher probability of infection [19–23]. Thus, centrality measures can identify individuals at high risk of infection and are often proposed as a method of guiding the distribution of vaccines and testing resources during outbreaks [7, 18, 24]. However, the predictive performance of centrality depends on the centrality measure used, the transmission mode of the parasite, whether the parasite is epidemic or endemic, and the social system being modeled. For example, one study found that centrality-based targeting of vaccines in response to an epidemic in wealthy countries appeared less effective than other targeting methods when parasites had a high *R_0_* and inter-regional travel rates were high [18].

On many social networks, parasites can spread through multiple transmission pathways. These different transmission pathways can involve different types of social relationships and processes, such as sexual contact versus needle sharing in the spread of HIV, or shared environmental exposures versus social contact in the spread of gastrointestinal parasites [23, 25–30]. These processes often involve different probabilities of transmission along network edges that can greatly influence disease dynamics. In the case of HIV, for example, needle sharing is associated with a higher risk of transmission than sexual intercourse, and among sexual contacts, transmission from males to females is higher than the reverse [25]. Thus, centrality on its own misses important dimensions of social relationships like the differential risk of transmission within and among groups and the social factors that drive transmission dynamics. Understanding these social roles can help guide public health interventions beyond simple estimates of superspreading potential based on single-relation centrality scores.

Prior network studies of infectious disease transmission have attempted to account for these differences in transmission probability. For example, to better understand the heterogeneous effects of non-pharmaceutical interventions during the COVID-19 pandemic, one study varied the relative edge weights of social contacts within and outside of the household and found that social distancing was most effective when within household contacts were weighted either very high or very low relative to external contacts [31]. Another approach is to create an abstracted measure of social activity that aggregates interaction information across a whole network. For example, social fluidity is a measure that accounts for both the density and weight of edges and can be used to assess the gregariousness of a social group and its effect on transmission across different domains of interaction [32].

A third approach – and the focus of this paper – is role analysis via equivalence-based blockmodeling. This approach clusters individuals who have similar patterns of social ties but who are not necessarily connected to the same people [33–36]. For any directed network, there are 36 positions an individual can occupy in a triad, and individuals are considered regularly equivalent if they occupy the same triad positions **(Figure 1)**. Similarity in centrality measures can also be used to provide additional information about shared patterns of social connections. The resulting roles thus reflect distinct patterns of *how* individuals are connected across networks – i.e., their types of relationships – rather than *whom* specifically they are connected to.

**Figure 1.**
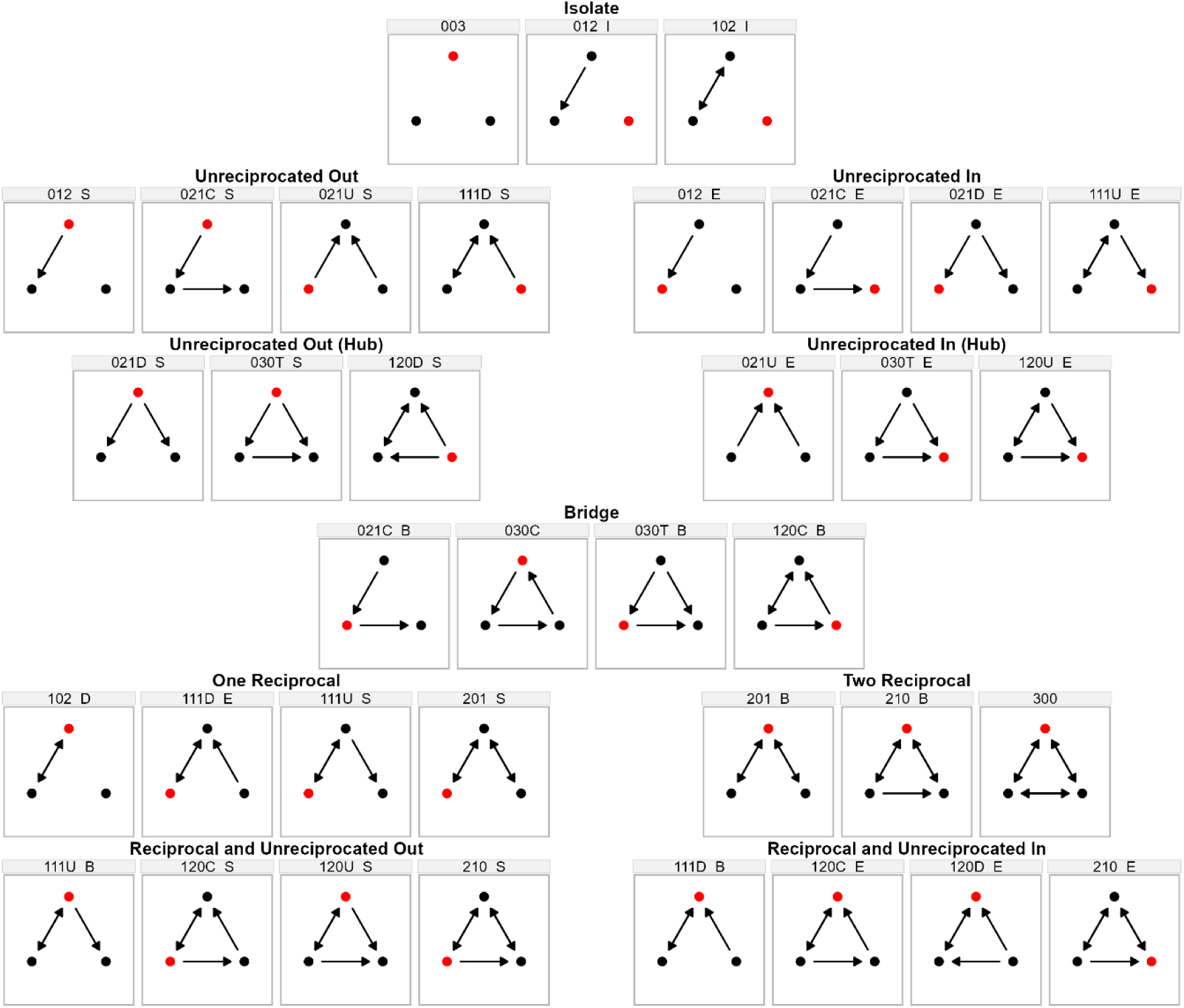
Typology of triad positions. Each panel shows one of 36 triad positions in 16 directed triads, with the focal node in red.

In infectious disease ecology and network epidemiology, role analysis can identify classes of nodes with equivalent risk profiles across multiple relations simultaneously. While traditional centrality scores used in epidemiology capture one dimension of transmission risk within one relation (e.g., degree being most relevant for local transmission and betweenness for network-wide transmission), assigning roles based on blockmodels allows a researcher to differentiate subsets of nodes with similar connection profiles and to identify how distinct profiles might contribute to different parasite transmission outcomes.

In the social sciences, role analysis has helped answer research questions about how informal social roles are related to marriage exchange, the accumulation of political power, political polarization, and the formation of social hierarchies [37–40]. To date, role analysis has had less influence in ecology and evolutionary biology, but it has been used to study food webs in Malaysia and Florida as well as the social structure of rhesus monkeys (*Macaca mulata*) in Pakistan and Cayo Santiago [41, 42]. To our knowledge, role analysis has not yet been applied to disease ecology or network epidemiology.

Here, we characterized distinct social roles based on friendship, food exchange, and farm co-working in three rural agricultural villages in northeast Madagascar using regular equivalence-based blockmodels. We then assessed whether these roles are associated with different patterns of viral exposure using a multiplex serological assay (Phage ImmunoPrecipitation Sequencing [PhIP-Seq]) with the VirScan library [43, 44]. PhIP-Seq VirScan captures the presence of antibodies to over 300 virus species and subtypes, allowing for the broad characterization of an individual’s history of viral exposure. Prior studies have used VirScan to determine differences in viral exposure profiles between Batwa hunter-gatherers and Bakiga agriculturalists in Uganda, finding that the hunter-gather population had relatively higher seropositivity of double-stranded DNA viruses [45]. Given the durability of social roles [46, 47], we expected the roles identified via equivalence-based blockmodels in rural Madagascar to reflect relatively long-term and distinct patterns of engaging in social life, which in turn might influence an individual’s viral encounters.

Our study had two primary objectives. First, we aimed to provide a case study of how regular equivalence blockmodeling can provide a richer characterization of a social system beyond individual centrality measures. Second, we aimed to assess whether the roles identified were epidemiologically relevant (hereafter, called social-epidemiological roles). Within this objective, we hypothesized that individuals who were most active and gregarious in their social interactions (i.e., had high centrality across networks and sent and received many different social ties) would be exposed to more viruses. We also expected that the social roles would perform better at predicting viral exposure than single measures of centrality. In testing our hypotheses, we accounted for sociodemographic characteristics that often affect infectious disease exposure, including age, gender, education, household size, and wealth.

## Methods

### Data Collection

Survey data were collected using snowball sampling from 1,297 adults (≥18 years old) in three villages (Village A, B, and C) bordering Marojejy National Park in the SAVA region of northeast Madagascar [48]. All data were collected between October 3, 2019 and August 4, 2022, and the study was approved by the Duke University IRB (protocol no. 2019-0560) and the Malagasy Ethics Committee for Biomedical Research within the Ministry of Public Health (114/MSANP/SG/AGMED/CERBM). All study participants provided informed consent and were compensated for their study participation.

Surveys were administered in the local Malagasy dialect by a trained research team to collect information about participants’ socioeconomic characteristics and social networks. An initial set of participants who owned agricultural land were identified in each village. We then used snowball sampling to recruit participants who were named as someone with whom a participant spent free time, exchanged food, or exchanged farmwork [49]. Each participant reported their age, gender, highest level of education, occupation, and household size.

We used two measures to assess participants’ market-based wealth [26]. First, we used an inventory of the durable goods an individual’s household owned, including whether their household owned a mobile phone, television, bicycle, motorcycle, generator, refrigerator, and computer [50, 51]. We then created an index with values ranging from 0 (no durable goods owned) to 7 (all durable goods owned). Second, we created an index of the material used to construct the floor, walls, and roof of a participant’s house, ranging from more gatherable materials (e.g., dirt, bamboo, raffia palm) to more expensive purchased materials (e.g., bricks, concrete, metal roofing sheets) [26, 52]. Each house component was ranked along this spectrum and then the values were *z*-score standardized. The standardized values for each house component were summed to create the final index (house lifestyle index) with higher values indicating greater market-based wealth and lower values indicating less market-based wealth.

We used five name-generating survey questions to construct the social networks used in this analysis, representing different types of relations [48]. Thus, all participants were asked to name up to five people with whom they spent their free time, provided and received food assistance, and provided and received farmwork assistance. Each of the five directed networks were included in the role analysis. To create single summary measures of centrality, we created a weighted network for each of the three villages where edge weights ranged from 0 (no types of relations) to 5 (all types of relations). In the summary networks, Village A (estimated village population size = 2,700) had 262 nodes and 952 edges, Village B (estimated village population size = 900) had 435 nodes and 1,642 edges, and Village C (estimated village population size = 1,900) had 600 nodes and 3,049 edges. These networks have been characterized in more detail in prior work [53].

### Phage ImmunoPreccipitation Sequencing

Dried blood spot samples were collected on Whatman 903 filter paper cards. Filter paper cards were stored at 0°C to −5°C in the field, followed by −5°C to −20°C at a storage site in Madagascar until they were shipped to Singapore and stored at −20°C to −80°C until analysis. Blood samples from the filter paper cards were extracted using a solution of 0.05% Tween-20 in Phosphate Buffered Saline (PBS). Punched out blood spots were incubated with the extraction solution overnight. Debris was then excluded by centrifugation, and the resulting supernatant was used for subsequent analysis.

PhIP-Seq VirScan was performed using previously described protocols [54, 55]. Briefly, the extracted samples were incubated with the VirScan bacteriophage library [44]. The resulting mixture was then immunoprecipited with Protein A and Protein G Dynabeads. A wash step was then performed, which was followed by nested PCR reactions to prepare the library for high throughput sequencing. This process extracts the gene of interest (coded in the bacteriophage genome) and appends a barcode sequence to allow for the multiplexing of samples in a sequencing run. High throughput sequencing was performed with BGI DNBSeq-G400 150-bp paired-end (BGI Hong Kong). Resulting sequences were demultiplexed and analyzed using in-house scripts described previously [54].

### Data Analysis

All data analyses were conducted with R version 4.4.0 [56]. To identify social roles, we used the role_analysis function in the *ideanet* package [57]. We operationalized roles using regular equivalence, which characterizes individuals who have similar patterns of social ties but who are not necessarily connected to the same people [58]. For each individual participant, we generated a vector of counts of the number of times that they occupied each of the 36 triad positions in each of the five networks [34]. We then augmented this triad position vector with centrality measures computed for each individual participant in each network. This augmentation included a wide range of transmission-relevant measures: total degree, indegree, outdegree, betweenness, power, eigenvector, and closeness centrality. Similarity in participants’ triad position and centrality vectors was determined using hierarchical clustering, and the final roles were determined by maximizing cluster modularity [36].

We then characterized the resulting blockmodel for each village and identified a typology of role categories across villages. To provide a high-level characterization of the role categories, we grouped the triad positions into four structural patterns, including reciprocal, unreciprocated out (senders), unreciprocated in (receivers), and bridging. For each village and role category combination, we calculated the mean frequency for triad positions reflecting each of these structural patterns and the mean values for centrality scores. The means were then *z*-score standardized within villages to compare relative triad position frequencies and centrality scores among role categories, and these *z*-scores were averaged across villages to produce a summary of relative triad positions and centrality scores across role categories.

We took two approaches to assess the relationship between social roles and viral exposure profiles. First, we assessed the relationship between role category and exposure to viruses that had >10% seroprevalence in the sample. To do so, we fit a Bayesian multilevel logistic regression model with weakly informative priors using the *rethinking* package [59]. The model included age, gender, education, occupation, household size, durable goods ownership, housing material, village, and role category as predictor variables and virus as a random effect. We computed pairwise differences in exposure probabilities between role categories by subtracting their posterior probability distributions.

Second, we assessed the importance of role category as a predictor of viral species and subtype richness (i.e., the number of virus species and subtypes an individual has antibodies to) using a multi-model comparison and averaging approach with the *MuMIn* package [60]. Each model used a zero-inflated Poisson distribution to account for zero-inflation in richness. We compared the predictive performance of models containing different sets of predictor variables (age, gender, education, occupation, household size, durable goods ownership, housing material, village, individual centrality measures [strength, betweenness, eigenvector, and closeness], and role category) using sample-size adjusted Akaike Information Criterion (AICc) [61, 62]. This approach allowed us to assess the direction of the relationship between role categories and richness. It also provided a means to compare the importance of role categories relative to demographic characteristics and individual centrality measures for predicting richness, with importance measured as the summed AICc for all models that included that variable. All continuous variables were standardized prior to the analysis, and weighted coefficients were computed across a set of models with a delta AICc less than two.

## Results

### Participant Demographic Characteristics

Participant demographic characteristics for each village are presented in **Table 1**. Overall, participants’ median (IQR) age was 33 (24-48) years, and 46% (n = 593) were women. Nearly all participants (96%, n = 1,237) had at least some education, and 12% (n = 158) had above a secondary-level education. Most (90%, n = 1,166) reported farming as their main occupation. Median (IQR) household size was 4 (3-5) people, and participants had a median (IQR) household lifestyle index of 0.63 (−1.55-1.84), indicating that their homes tended to be constructed of purchased materials (e.g., metal or concrete) rather than materials readily available in the environment (e.g., wood or ravinala palm). Participants reported that their household owned a median (IQR) of 1 (0-2) durable goods.

**Table 1.** Participant demographic characteristics by village.

| **Demographic Characteristics** | **Village A**<br>**(n = 262)** | **Village B**<br>**(n = 435)** | **Village C**<br>**(n = 600)** | **All Villages**<br>**(n = 1,297)** |
| --- | --- | --- | --- | --- |
| Age, median (IQR) | 40 (30-53) | 30 (22-44) | 32 (24-47) | 33 (24-48) |
| Unknown | 1 | 1 | 0 | 2 |
| Gender, n (%) |  |  |  |  |
| Women | 120 (46%) | 182 (42%) | 291 (49%) | 593 (46%) |
| Men | 142 (54%) | 253 (58%) | 309 (52%) | 704 (54%) |
| Education, n (%) |  |  |  |  |
| None | 14 (5.3%) | 21 (4.9%) | 14 (2.4%) | 49 (3.8%) |
| Primary | 158 (60%) | 179 (41%) | 243 (41%) | 580 (45%) |
| Secondary | 67 (26%) | 178 (41%) | 254 (43%) | 499 (39%) |
| Higher | 23 (8.8%) | 54 (13%) | 81 (14%) | 158 (12%) |
| Unknown | 8 (3%) | 0 (0%) | 3 (0.5%) | 11 (0.8%) |
| Occupation, n (%) |  |  |  |  |
| Farmer | 256 (98%) | 388 (89%) | 522 (87%) | 1,166 (90%) |
| Non-Farmer<sup>1</sup> | 6 (2.3%) | 46 (11%) | 76 (13 %) | 128 (9.9%) |

| Unknown | 0 (0%) | 1 (0.2%) | 2 (0.3%) | 3 (0.2%) |
| --- | --- | --- | --- | --- |
| House Lifestyle Index, median (IQR) | -1.55 (-1.55-1.84) | 0.63 (-0.34-1.84) | 0.63 (-1.55-1.84) | 0.63 (-1.55-1.84) |
| Durable Goods Owned, median (IQR) | 1 (0-2) | 1 (0-2) | 1 (0-2) | 1 (0-2) |
| Household Size, median (IQR) | 4 (3-5) | 4 (3-5) | 4 (3-5) | 4 (3-5) |

<sup>1</sup>Non-farming occupations included carpenter, taxi driver, laborer, shopkeeper, teacher, student, and unemployed.

### Role Analysis

Eight distinct clusters were identified for Village A. Five clusters were identified for Village B, and four clusters were identified for Village C **(Figure 2)**. The role structure across all three villages suggested three primary categories of roles: (1) individuals who were “Popular” and named by many individuals across nearly all roles in a village, (2) individuals who were “Hangers-On” and relatively central to a village network but who mostly named others in the village rather than being named themselves, and (3) individuals on the “Periphery” of a village network and minimally connected to the Popular individuals. Age, gender, education, and occupation were relatively similar across the three role categories **(Table 2)**.

**Figure 2.**
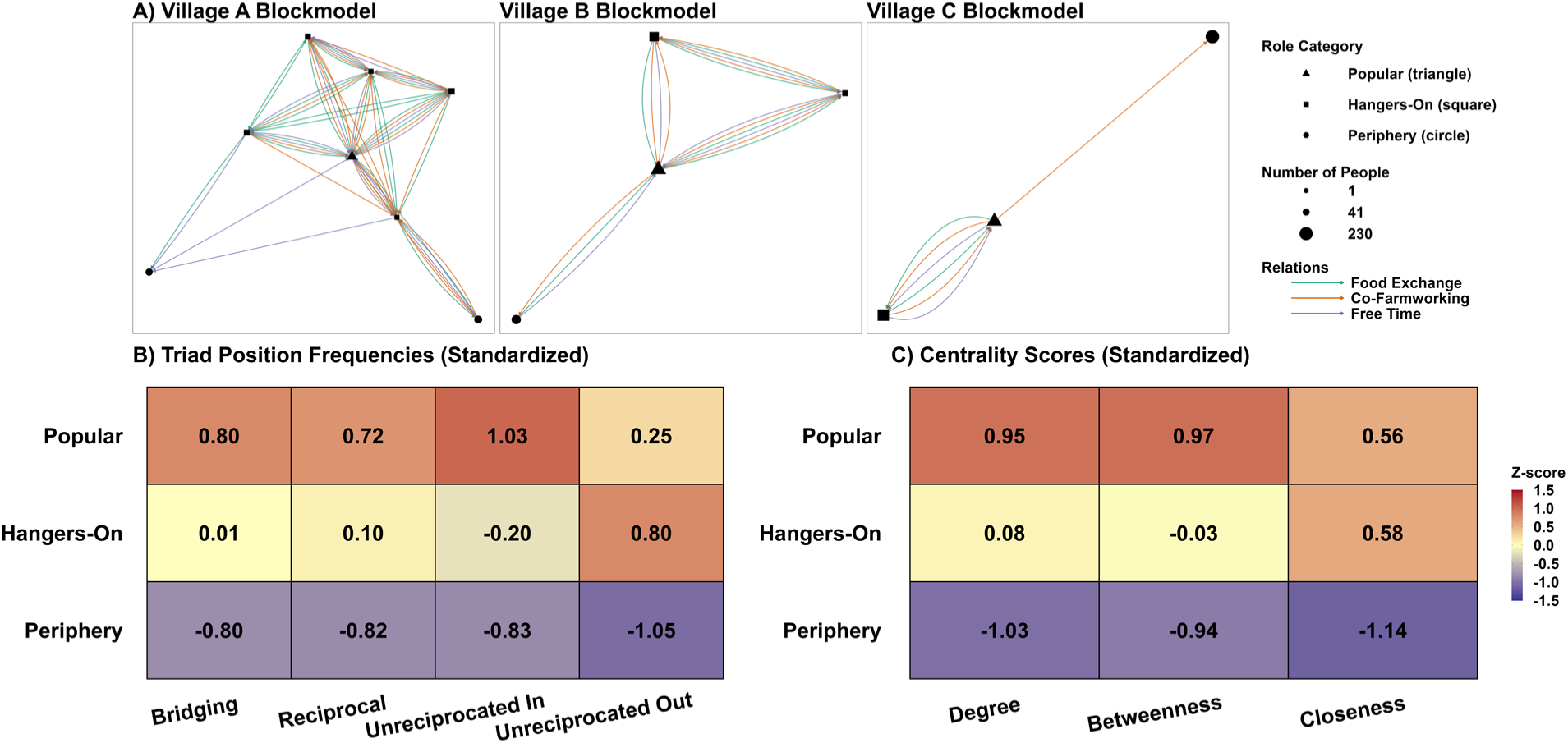
Role analysis summary. Panel A shows the blockmodels for the three villages, with nodes representing roles in the Popular (triangle), Hangers-On (square), and Periphery (circle) role categories. Edges between nodes indicate whether members of one role nominated members of another role as people with whom they spend free time (purple), exchange food (green), and co-farmwork (orange). Panels B and C highlight a selection of key characteristics for each role category. Panel B shows the standardized frequency at which members of a role category occupied different types of triad positions averaged across relations and villages. Panel C shows the standardized centrality scores for members of each role category averaged across relations and villages.

**Table 2.** Participant demographic characteristics by role category.

| Demographic Characteristics | Periphery<br>(n = 445) | Hangers-On<br>(n = 476) | Popular<br>(n = 376) | All Roles<br>(n = 1,297) |
| --- | --- | --- | --- | --- |
| Age, median (IQR) | 32 (22-46) | 33 (24-48) | 35 (26-49) | 33 (24-48) |
| Unknown | 1 | 0 | 1 | 2 |
| Gender, n (%) |  |  |  |  |
| Women | 209 (47%) | 220 (46%) | 164 (44%) | 593 (46%) |
| Men | 236 (53%) | 256 (54%) | 212 (56%) | 704 (54%) |
| Education, n (%) |  |  |  |  |
| None | 22 (5%) | 18 (3.8%) | 9 (2.4%) | 49 (3.8%) |
| Primary | 193 (44%) | 217 (46%) | 170 (45%) | 580 (45%) |
| Secondary | 169 (38%) | 178 (38%) | 152 (41%) | 499 (39%) |
| Higher | 55 (13%) | 59 (13%) | 44 (12%) | 158 (12%) |
| Unknown | 6 | 4 | 1 | 11 (0.8%) |
| Occupation, n (%) |  |  |  |  |
| Farmer | 389 (88%) | 427 (90%) | 350 (93%) | 1,166 (90%) |
| Non-Farmer<sup>1</sup> | 55 (12%) | 47 (9.9%) | 26 (6.9%) | 128 (9.9%) |

| Unknown | 1 | 2 | 0 | 3 (0.2%) |
| --- | --- | --- | --- | --- |
| House Lifestyle Index, median (IQR) | -0.34 (-1.55-1.84) | 0.63 (-1.55-1.84) | 0.63 (-1.55-1.84) | 0.63 (-1.55-1.84) |
| Durable Goods Owned, median (IQR) | 1 (0-2) | 1 (0-2) | 1 (1-2) | 1 (0-2) |
| Household Size, median (IQR) | 4 (3-5) | 4 (3-5) | 4 (3-5) | 4 (3-5) |

<sup>1</sup>Non-farming occupations included carpenter, taxi driver, laborer, shopkeeper, teacher, student, and unemployed.

Individuals in the Popular role category occupied reciprocal and unreciprocated-in triad positions at a higher frequency than the other role categories, indicating that they rarely named someone who did not also name them. They also occupied more bridging triad positions and had higher betweenness centrality than the other role categories. Together with their relatively high degree centrality, these patterns suggest that members of the Popular role category were top of mind when participants were asked with whom they spend free time, exchange food, and participate in co-farmworking. These patterns also suggest that these participants’ connections spanned social groups within the villages.

Members of the Hangers-On role category occupied bridging, reciprocal, and unreciprocated-in triad positions at a comparatively lower frequency. Instead, they occupied unreciprocated-out triad positions at a much higher frequency. This pattern indicates that they named many other people in the network, but few people named them in return. They had closeness centrality scores comparable to members of the Popular role category, which suggests they were typically a short path away from other nodes in the network. In part, their higher closeness centrality is because they frequently named individuals in the Popular role category.

In general, members of the Periphery role category were represented in each type of triad position at a lower frequency than members of both the Hangers-On and Popular role categories. They also had much lower degree, betweenness, and closeness centrality scores. This pattern suggests that they engaged in exchange with a few highly influential individuals in each village, which likely brought them into the networks despite their minimal engagement with most members of a village. Standardized triad position frequencies and centrality scores for each village, relation, and role category are shown in **Figures S1-S6**.

### Association Between Role Categories and Viral Exposure

For a subset of participants from Village B and Village C with VirScan data (n = 156), we assessed the relationship between role categories and past viral exposure. Of the 337 virus species and subtypes in the VirScan library, antibodies to 62 (18%) of the virus species and subtypes were identified in these participants **(Figure 3)**. Ten of the 62 virus species and subtypes (16%) had a seroprevalence greater than 10%, including vaccina virus, orf virus, molluscum contagiosum virus subtype 1, influenza A virus, human respiratory syncytial virus, human herpesvirus 8, human herpesvirus 1, human cytomegalovirus, hepatitis B, and Epstein-Barr virus.

**Figure 3.**
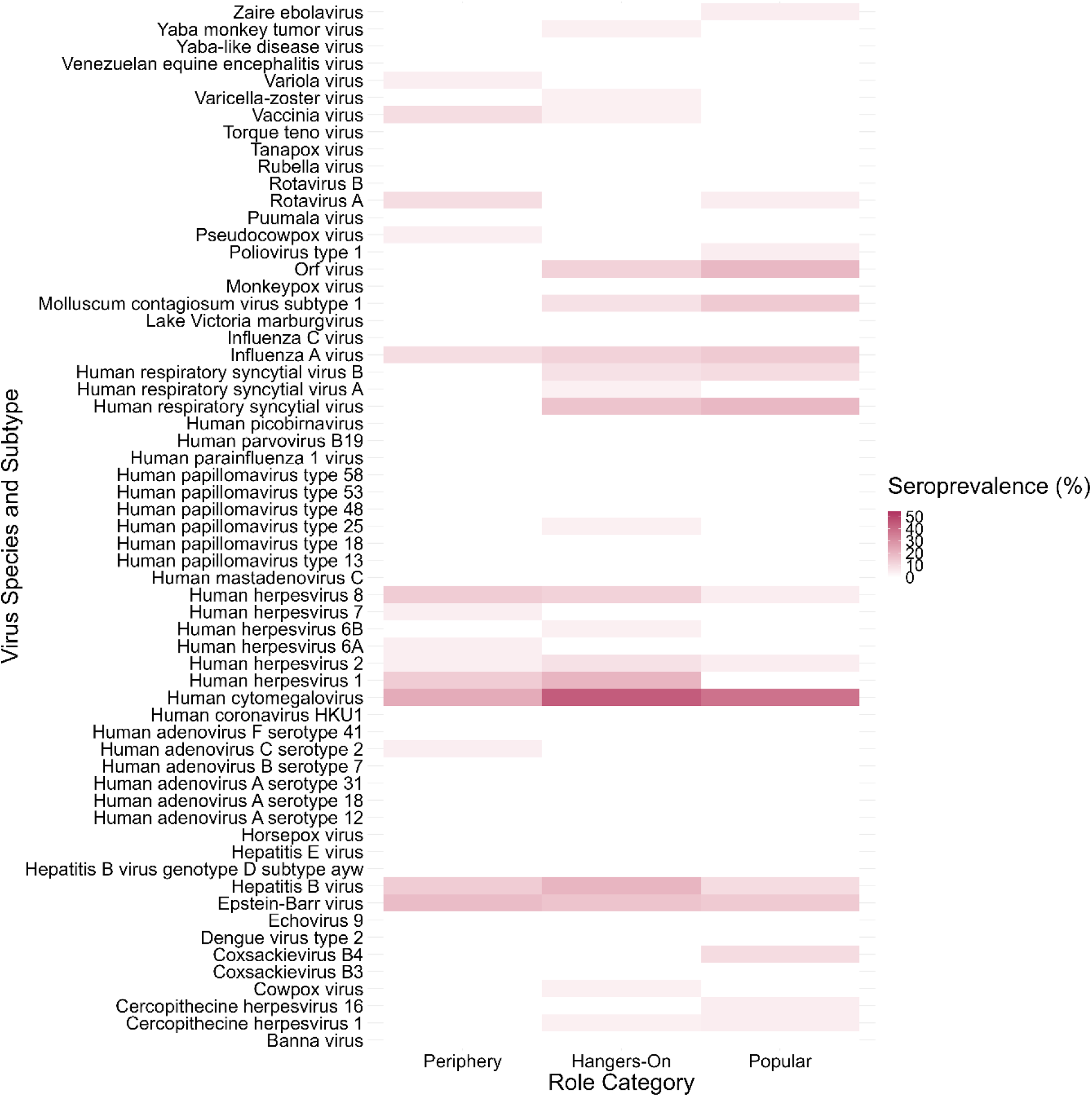
Seroprevalence by role category for the 62 virus species and subtypes identified in the sample. Darker red tiles indicate higher seroprevalence.

In a Bayesian multilevel logistic regression model predicting exposure to any of the ten common viruses, individuals in the Hangers-On role category had elevated exposure risk relative to both the Periphery (mean difference [95% CI]: 0.09 [0.03-0.15]) and Popular role categories (mean difference [95% CI]: 0.09 [0.03-0.17]) **(Figure 4)**. In contrast, the Popular and Periphery role categories did not meaningfully differ in their exposure probability (mean difference [95% CI]: −0.01 [−0.08-0.07]). Together, these results indicated that individuals in the Hangers-On role category had an approximately 8-9% higher exposure probability compared to the Popular and Periphery role categories, while accounting for sociodemographic characteristics. The complete model results are reported in **Table S1**.

**Figure 4.**
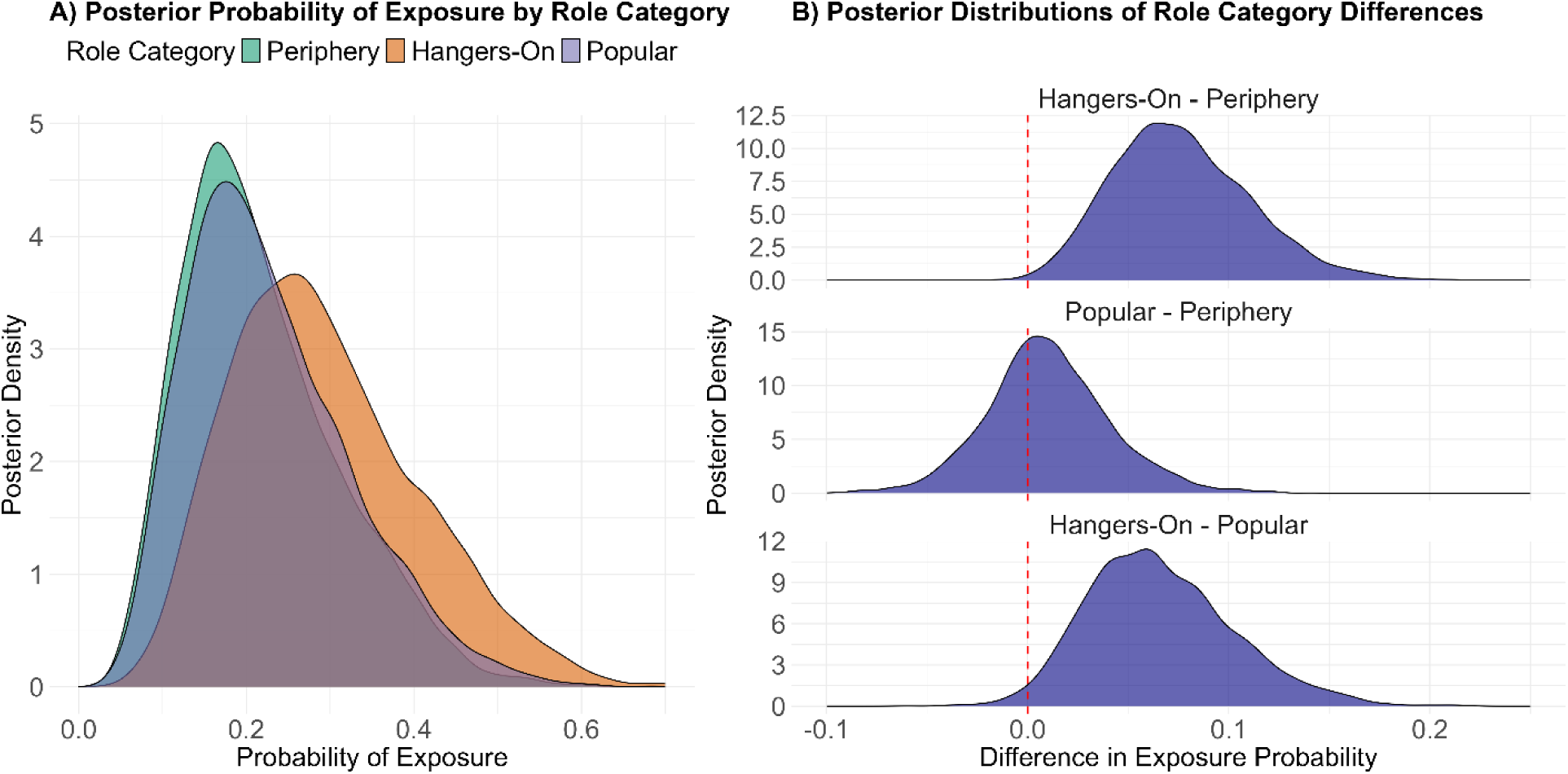
Probability of viral exposure by role category. Panel A shows the posterior probability of exposure by role category to at least one of ten viruses with greater than 10% seroprevalence. The model also included participant sociodemographic characteristics as covariates and virus as random effect. Panel B shows the difference in exposure probability between each of the three role categories.

Participants had a median (IQR) viral species and subtype richness of 2 (1-4). Role category, education, and village were the most important predictors of richness (summed AICc weight = 1; **Figure 5**). Membership in both the Hangers-On role category (model averaged *ß* [95% CI]: 0.36 [0.12-0.60]) and Popular role category (model averaged *ß* [95% CI]: 0.29 [−0.06-0.63]) was associated with higher richness than membership in the Periphery role category. Residence in Village B was associated with lower richness than residence in Village C (model averaged *ß* [95% CI]: −0.79 [−1.07-−0.52]). Eigenvector centrality was a relatively important predictor of richness (summed AICc weight = 0.64), but all four individual centrality measures had a coefficient close to zero and were less important than the role categories. Strength (summed AICc weight = 0.15) and closeness centrality (summed AICc weight = 0.07) were both of minimal importance for predicting richness, and betweenness centrality was not included in any of the eleven component models used in the model averaging **(Table S2)**. Age, gender, and the two measures of market-based wealth all had a summed AICc weights less than 0.2, and the direction of each relationship was unclear.

**Figure 5.**
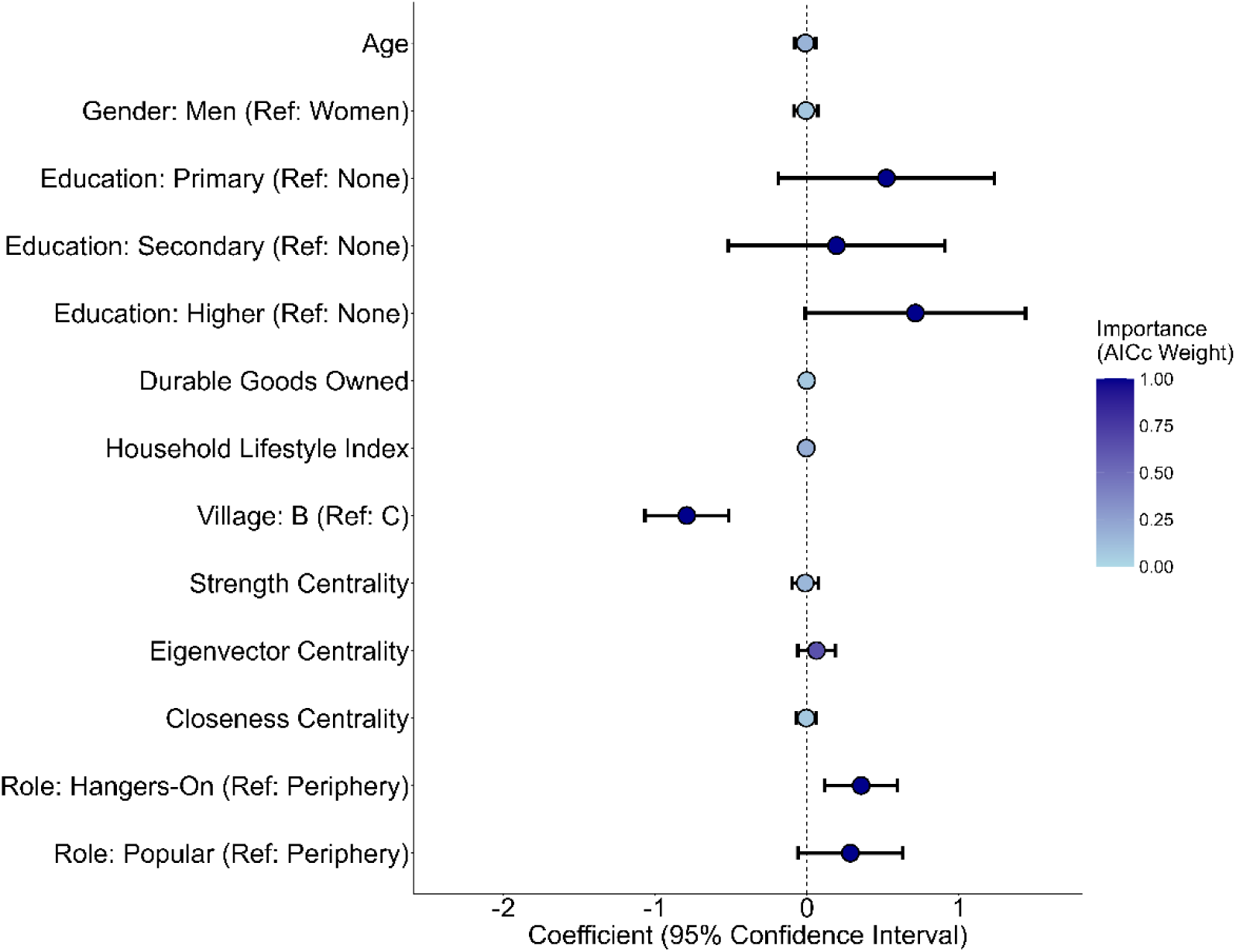
Association between predictor variables and virus species and subtype richness. Points and error bars represent *ß* coefficients and 95% confidence intervals from model averaging, and darker blue points reflect variables that were more important (higher AICc weight) for predicting viral species richness.

## Discussion

Measures of node centrality can provide important information for predicting which individuals in a social network are most likely to be exposed to a parasite and have the greatest spreading potential [6–12]. In this study, we demonstrated that the utility of single centrality measures is enhanced by clustering individuals by their similarity in both centrality scores and triad positions across multiple networks or types of relations. We used this approach – role analysis via equivalence-based blockmodeling – to identify social-epidemiological roles associated with viral exposure in three rural villages in northeast Madagascar. We identified three role categories (Popular, Hangers-On, and Periphery) that reflected general patterns in how people tended to spend their free time with others and exchange food and farmwork. We found partial support for our hypothesis that individuals with high centrality scores who named and were named by many others would be exposed to the most viruses. Specifically, we found that membership in the Hangers-On role category – those who sent many ties with few reciprocated – had the strongest positive association with viral exposure. Members of the Hangers-On role category also had relatively high closeness centrality, indicating that they were a short distance from everyone else in the network. As expected, we found that social-epidemiological roles were more predictive of viral exposure than measures of centrality, suggesting that role analysis via equivalence-based blockmodeling can capture important information about how people interact across relations in ways that are meaningful for understanding infectious disease transmission dynamics.

The role categories identified in our analysis provided distinct insights into viral transmission dynamics in a rural village setting. An individual’s vector of triad positions provides a summary of how they interact across different domains of social life [36], specifically how individuals socialize in their free time, how they help and are helped by others when food is scarce, and how they engage in farmwork with others. Given that greater social activity provides more opportunities for infectious disease exposure, we hypothesized that people who engaged in reciprocal relationships across these domains would be more socially active and thus have more opportunities for viral exposure. However, we found that membership in the Hangers-On role category – representing individuals who sent many ties across relations that were rarely reciprocated and who participated in few reciprocal closed triads – was associated with greater viral richness.

This finding suggests a potential alternative hypothesis. Sociality has both costs and benefits for infectious disease risk, with the primary cost being increased probability of exposure [63]. Low social status and chronic psychosocial stress can also weaken the immune system [64, 65]. Individuals who reported more unreciprocated relationships may thus experience stress-induced immunosuppression, and their high social activity – sending many ties – combined with lower social status – relative to members of the Popular role category – may make them particularly susceptible to infection. Our finding that individuals in the Hangers-On role category had the highest exposure risk for viruses with greater than 10% seroprevalence may also point to this hypothesis. Four of the viruses included in this group were herpesviruses. Prior studies linking social stressors to circulating antibody levels for herpesviruses like Epstein-Barr virus and cytomegalovirus have highlighted the importance of psychosocial stress in affecting susceptibility to infection and reactivation [66–71]. Further work is needed to more directly identify the mechanisms by which the social-epidemiological roles identified in this study may affect susceptibility to infectious diseases through stress pathways.

This study also revealed important differences in how measures of centrality relate to viral exposure in a rural village setting. Each role category had distinct patterns of centrality scores, but the role categories themselves were more important for predicting viral species and subtype richness than single measures of centrality alone. This finding suggests that role analysis via equivalence-based blockmodeling allows for a more complete representation of the social processes that underlie infectious disease transmission dynamics. For example, members of the Hangers-On role category had relatively high closeness centrality scores, and membership in this role category was associated with greater viral species and subtype richness. However, closeness centrality performed relatively poorly at predicting richness. Our findings suggest that the most at-risk individuals in a network might not simply be those who are most central but rather those who fit a general profile of being both central (increasing the probability of exposure) and lower social status (increasing susceptibility to infection once exposed).

In network epidemiology, approaches to identify the most at-risk individuals with high spreading potential often emphasize measures of centrality [6]. Our findings suggest that additional positional network features like an individual’s level of engagement in reciprocal relationships provide important information for understanding disease transmission and targeting interventions. The real-world application of this approach is limited by the need for network data, which are often limited or unavailable in low-resource settings [7]. However, general principles can emerge that guide interventions, and in cases where network data are available to compute centrality scores, no additional data are needed to perform regular equivalence blockmodeling. Our results suggest that performing this analysis provides added value. This approach may be particularly useful in disease ecology, where researchers are increasingly collecting data on multiple types of relations [26–28, 30]. Our study demonstrates how role analysis can be used in combination with these multidimensional datasets to uncover complex transmission dynamics and generate new transmission-relevant hypotheses.

Our study had several limitations. First, although we had comprehensive network data for all three study villages, our analysis of viral exposure was limited to a subset of participants from two of the three villages. This data limitation precluded an analysis of viral exposure in Village A, which had the most complex set of roles. Sample size limitations also required us to analyze roles at a higher level (role category) for Villages B and C, but the more granular roles could be used in studies with more data per role. Second, PhIP-Seq VirScan is a powerful tool for characterizing exposure to a broad set of viruses [43, 44]. However, determining seropositivity for a specific virus requires confirmatory analyses through more traditional methods like enzyme-linked immunosorbent assays (ELISAs). Third, we examined the utility of role analysis for predicting viral exposure, but its utility for other types of parasites might differ. Future work should explore applications across broad categories of parasites, from viruses to helminths.

## Conclusions

Infectious diseases often spread through multiple pathways, and analytical techniques for identifying high risk individuals in transmission networks can be limited by an overemphasis on single measures of centrality. We demonstrated the utility of role analysis via equivalence-based blockmodeling for identifying social-epidemiological roles that are associated distinct patterns of exposure to viruses in a rural village setting. The roles we identified performed better at predicting viral exposure than single measures of centrality alone, and the analysis informed new hypotheses for the coupled dynamics of susceptibility and exposure in viral transmission. Future studies in disease ecology and network epidemiology could benefit from using role analysis via equivalence-based blockmodeling to summarize increasingly high dimensional network data.

## Data Availability

The data that support the findings of this study are available on request from the corresponding author. The data are not publicly available due to privacy or ethical restrictions.

## Acknowledgements

We thank the Duke Lemur Center-SAVA Conservation for logistical support and the Malagasy Ethics Panel for permission to conduct the research. We greatly appreciate the three study communities for their participation and hospitality. Funding was provided by the joint NIH-NSF-NIFA Ecology and Evolution of Infectious Disease Program (R01-TW011493-01, DEB-2308460), an NSF Doctoral Dissertation Research Improvement Grant (BCS-2341234), an NSF Predictive Intelligence for Pandemic Prevention (PIPP) Phase I grant (SBE-2200047), and a Duke Provost’s Collaboratory grant.

