## Supplemental Material for "Identifying Social-Epidemiological Roles Associated with Viral Exposure Using Regular Equivalence Blockmodeling"

**Table S1.** Bayesian multilevel logistic regression model predicting exposure to any of ten viruses with seroprevalence > 10%. Virus species/subtype was included as a random effect.

| Predictor | Posterior Mean | 95% Credible Interval |
|---|---|---|
| Age | 0.08 | -0.05-0.20 |
| Gender | | |
| Men (Ref: Women) | -0.06 | -0.60-0.49 |
| Education | | |
| Primary (Ref: None) | -0.34 | -0.93-0.24 |
| Secondary (Ref: None) | 0.11 | -0.33-0.56 |
| Higher (Ref: None) | -0.18 | -0.61-0.28 |
| Household Size | 0.17 | 0.04-0.29 |
| Occupation | | |
| Non-Farmer (Ref: Farmer) | 0.07 | -0.49-0.63 |
| House Lifestyle Index | -0.16 | -0.27--0.05 |
| Durable Goods Owned | -0.06 | -0.19-0.07 |
| Role Category | | |
| Hangers-On (Ref: Periphery) | 0.43 | 0.18-0.67 |
| Popular (Ref: Periphery) | 0.06 | -0.24-0.35 |

**Table S2.** Component models predicting viral species and subtype richness that met the inclusion criteria for model averaging (delta AICc < 2)

| Model | Degrees of Freedom | Log Likelihood | AICc | Delta AICc | Weight | Age | Gender | Education | Occupation | Village | House Lifestyle Index | Durable Goods Owned | Strength | Betweenness | Eigenvector | Closeness | Role |
|---|---|---|---|---|---|---|---|---|---|---|---|---|---|---|---|---|---|
| 1 | 9 | -333.90 | 687.03 | 0.00 | 0.15 | | | ■ | | ■ | | | | | ■ | | ■ |
| 2 | 8 | -335.29 | 687.58 | 0.54 | 0.12 | | | ■ | | ■ | | | | | | | ■ |
| 3 | 10 | -333.22 | 687.96 | 0.93 | 0.10 | | | ■ | | ■ | ■ | | | | ■ | | ■ |
| 4 | 9 | -334.40 | 688.04 | 1.01 | 0.09 | | | ■ | | ■ | ■ | | | | | | ■ |
| 5 | 10 | -333.35 | 688.22 | 1.19 | 0.09 | ■ | | ■ | | ■ | | | | | ■ | | ■ |
| 6 | 9 | -334.51 | 688.27 | 1.23 | 0.08 | ■ | | ■ | | ■ | | | | | | | ■ |
| 7 | 10 | -333.40 | 688.33 | 1.29 | 0.08 | | | ■ | | ■ | | | ■ | | ■ | | ■ |
| 8 | 10 | -333.43 | 688.39 | 1.36 | 0.08 | | ■ | ■ | | ■ | | | | | ■ | | ■ |
| 9 | 10 | -333.50 | 688.54 | 1.51 | 0.07 | | | ■ | | ■ | | ■ | | | ■ | | ■ |
| 10 | 10 | -333.53 | 688.59 | 1.56 | 0.07 | | | ■ | | ■ | | | | | ■ | ■ | ■ |
| 11 | 9 | -334.77 | 688.78 | 1.75 | 0.06 | | | ■ | | ■ | | | ■ | | | | ■ |
| | | **Importance (Summed AICc Weight)** | | | | 0.17 | 0.08 | 1 | 0 | 1 | 0.19 | 0.07 | 0.15 | 0 | 0.64 | 0.07 | 1 |

**Figure S1.** Standardized triad position frequencies for Village A.

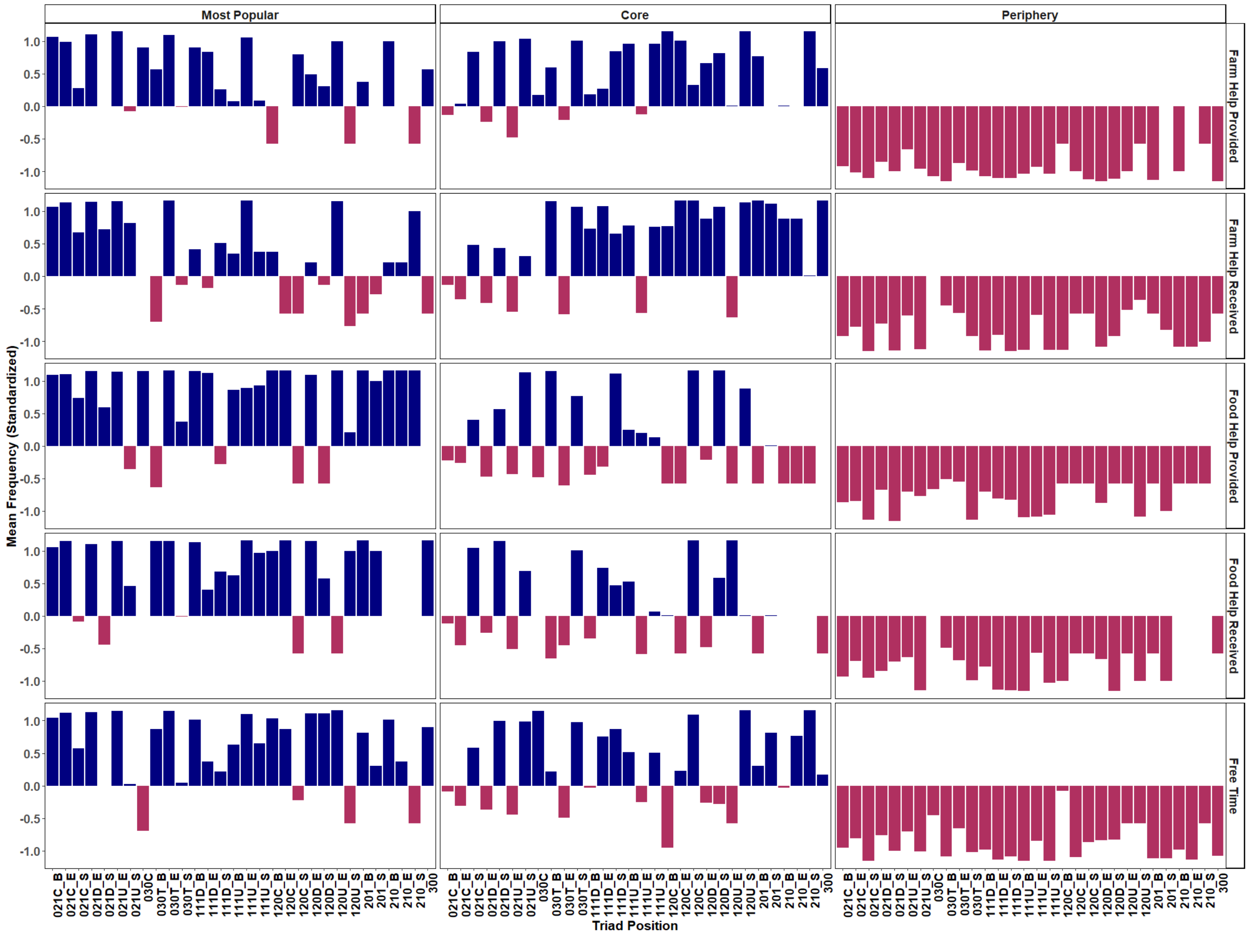

**Figure S2.** Standardized triad position frequencies for Village B.

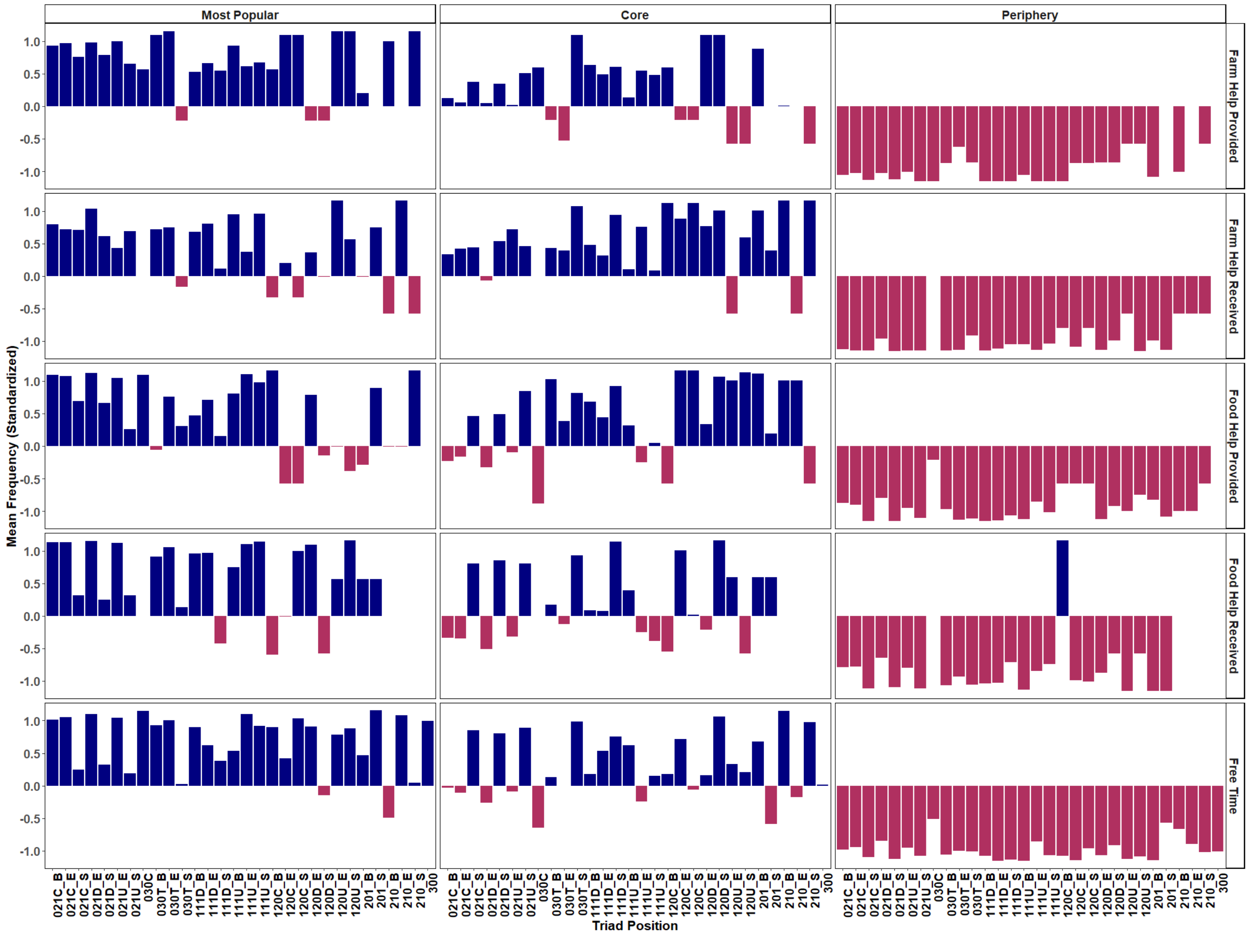

**Figure S3.** Standardized triad position frequencies for Village C.

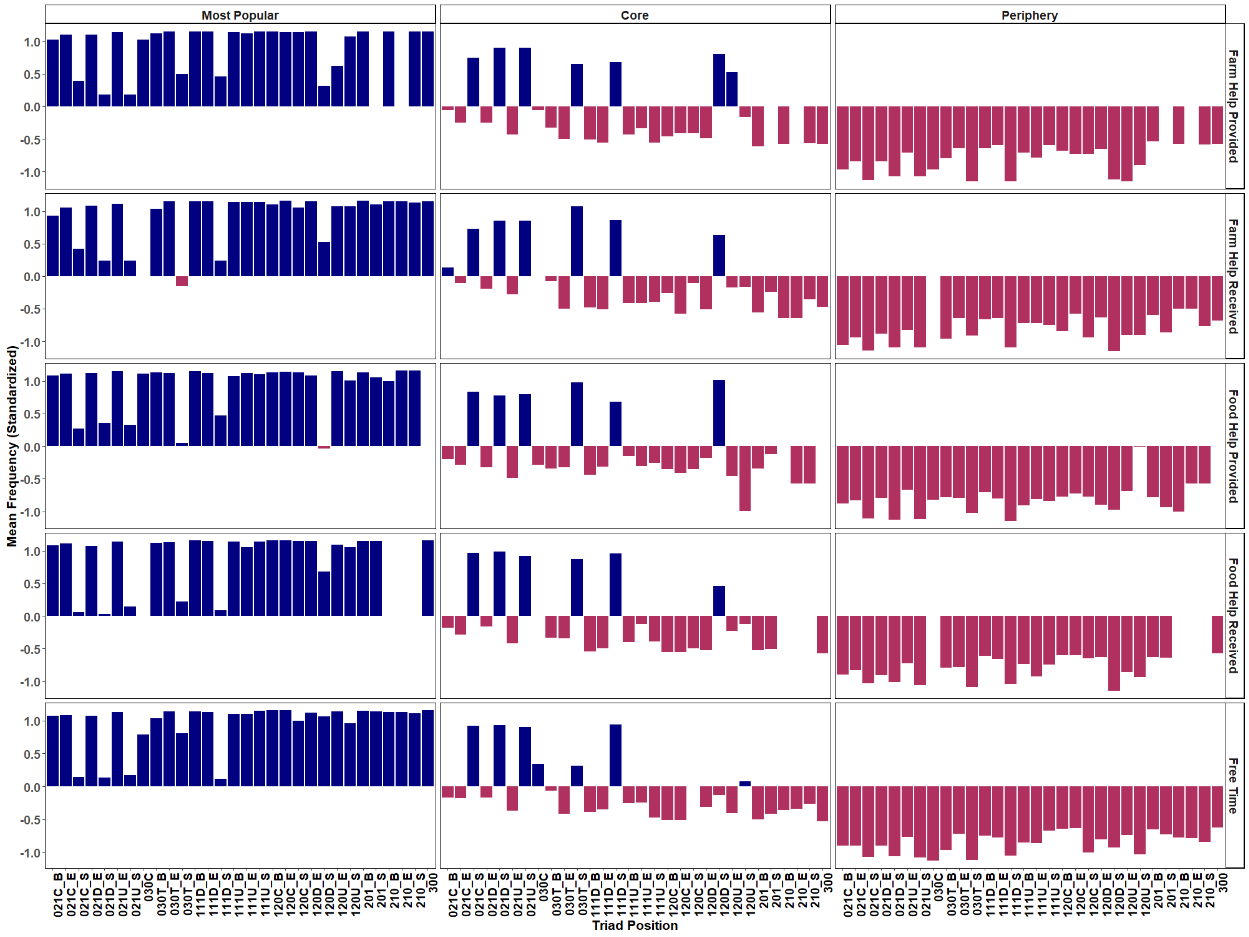

**Figure S4.** Standardized centrality scores for Village A.

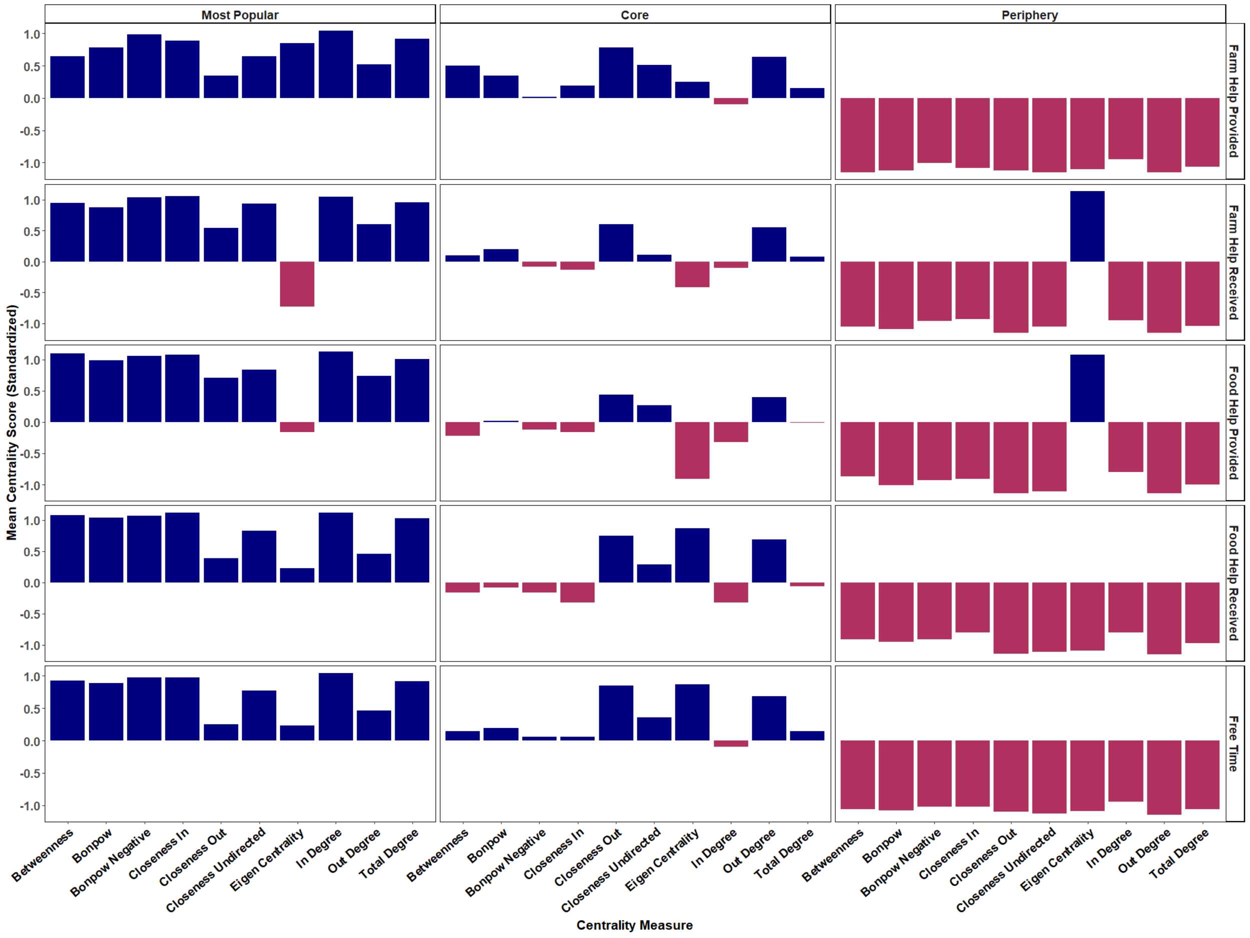

**Figure S5.** Standardized centrality scores for Village B.

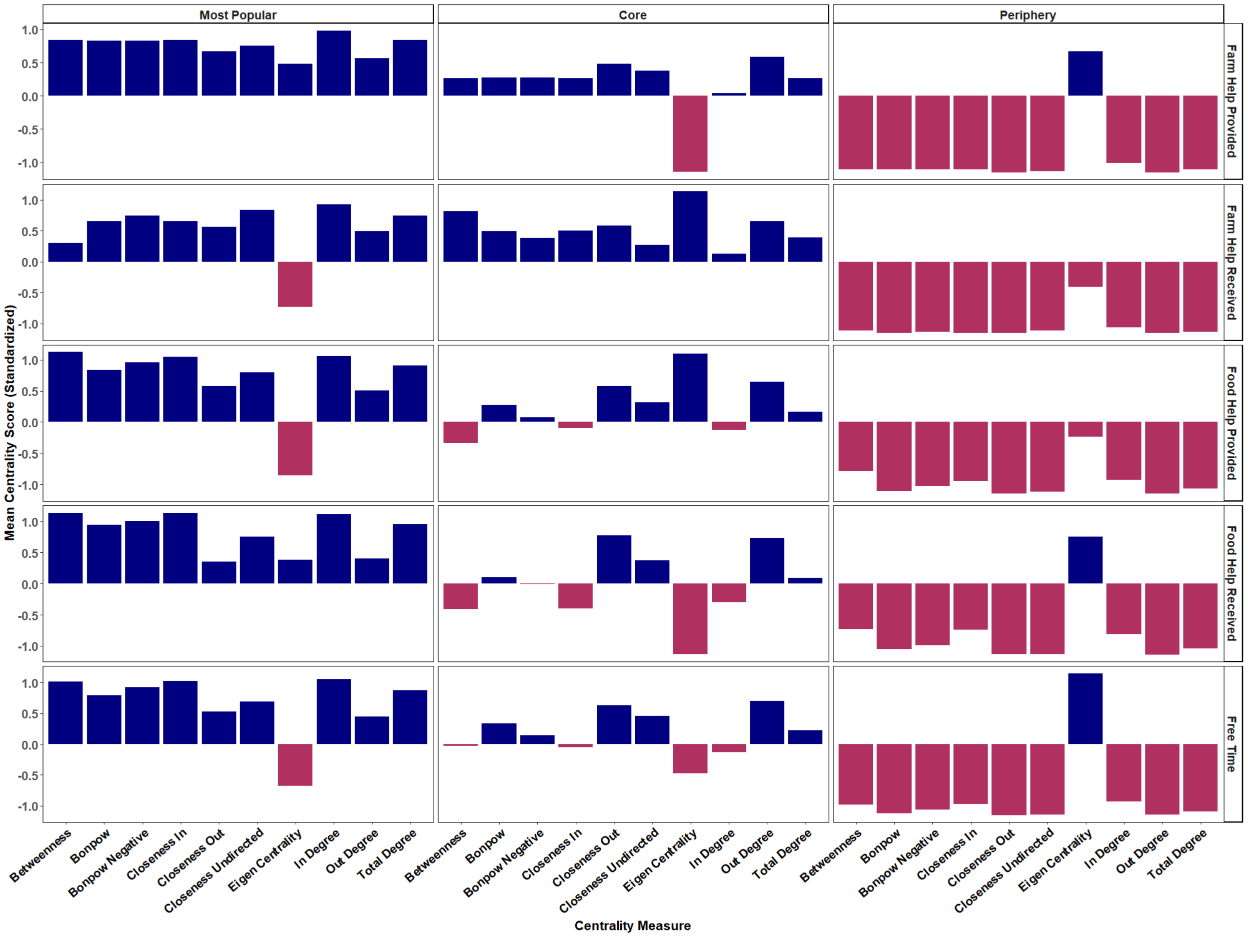

**Figure S6.** Standardized centrality scores for Village C.

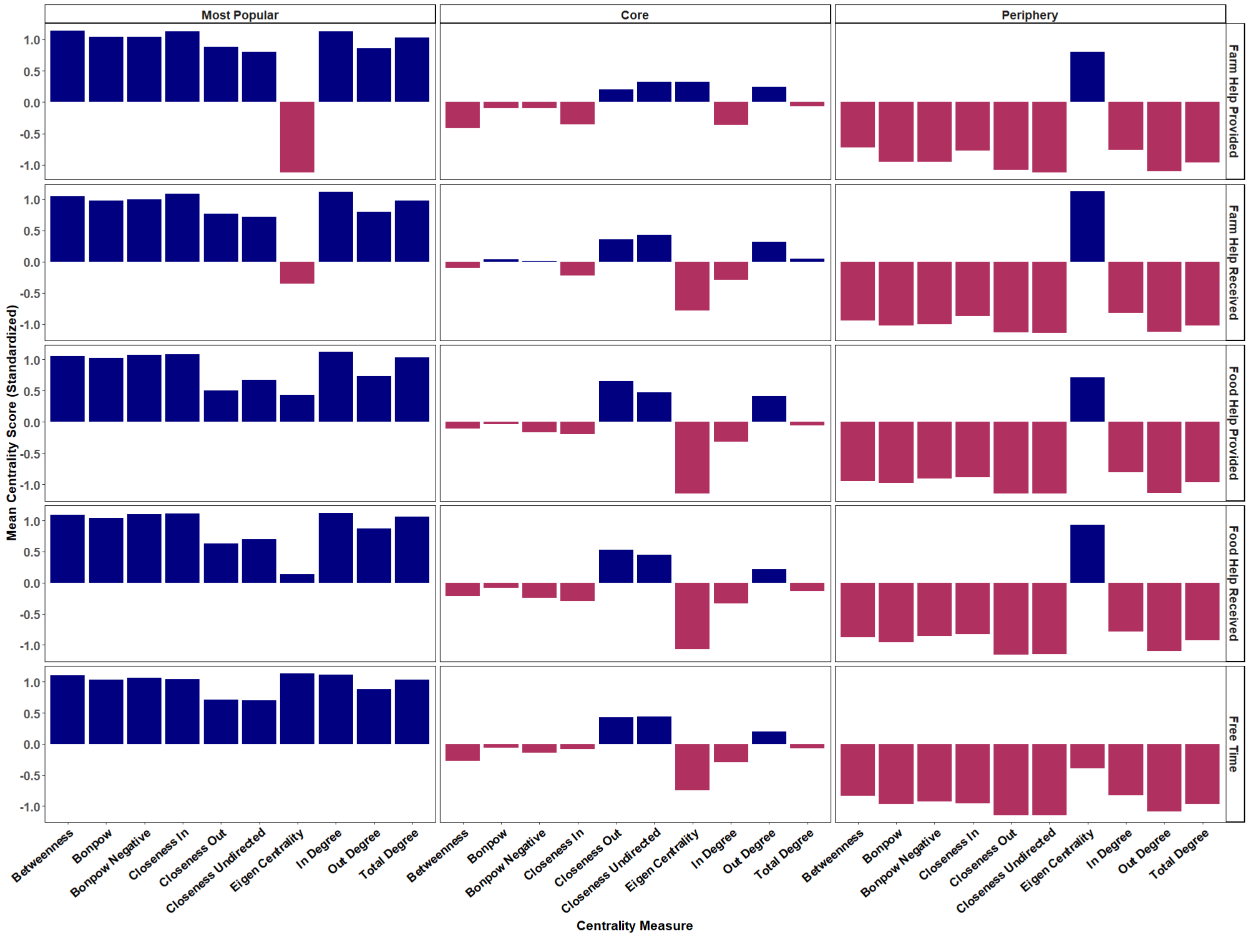
